# A Machine Learning Explanation of Incidence Inequalities of SARS-CoV-2 Across 88 Days in 157 Countries

**DOI:** 10.1101/2020.06.06.20124529

**Authors:** Eric Luellen

## Abstract

Because the SARS-CoV-2 (COVID-19) pandemic viral outbreaks will likely continue until effective vaccines are widely administered, (*1*) new capabilities to accurately predict incidence rates by location and time to know in advance the disease burden and specific needs for any given population are valuable to minimize morbidity and mortality. In this study, a random forest of 9,250 regression trees was applied to 6,941 observations of 13 statistically significant predictor variables targeting SARS-CoV-2 incidence rates per 100,000 across 88 days in 157 countries. One key finding is an algorithm that can predict the incidence rate per day of a SARS-CoV-2 epidemic cycle with a pseudo-R2 accuracy of 98.5% and explain 97.4% of the variances. Another key finding is the relative importance of 13 demographic, economic, environmental, and public health modulators to the SARS-CoV-2 incidence rate. Four factors proposed in earlier research as potential modulators have no statistically significant relationship with incidence rates (*2*)(*3*). These findings give leaders new capabilities for improved capacity planning and targeting stay-at-home interventions and prioritizing programming by knowing the atypical social determinants that are the root causes of SARS-CoV-2 incidence variance. This work also proves that machine learning can accurately and quickly explain disease dynamics for zoonoses with pandemic potential.

## Methods

In this study, machine learning – a robust statistical version of artificial intelligence – was applied to a data set of 6,941 observations to identify the relative importance of 13 demographic, economic, environmental, and public health factors in modulating the incidence rate per 100,000 population of SARS-CoV-2 across 88 days in 157 countries. The data was sourced from the public domain, such as the World Bank and United Nations, select journal articles, and weather stations. (*4*) The period of the epidemic curve measured began the day after cases in a country began to grow until case growth stopped or the 88-day period expired between January 23 and April 18, 2020.

Specifically, the author used the Rattle library (version 5.3.0; Togaware) in the programming language R (version 3.6.2, CRAN) to apply generalized linear models to learn the P-values of each term. From which four terms believed to be potential modulators of incidence rate were excluded because of P-values in excess of .05: maximum ultraviolet (UV) index (P-value = .348), minimum temperature (P-value = .896), humidity (P-value = .956), dengue fever incidence rate (P-value = .131), and median age (P-value = .062. Where after, the author applied a series of differently sized random forest algorithms, ranging from 500 to 10,000 regression trees, to learn the optimum number of regression trees to minimize error. The lowest error rate was approximately 9,250 regression trees, which the author applied, using four variables at a time, which was the closest whole number to the square root of the number of predictors.

The algorithm randomly partitioned the data to select and train on 70% (n=4858), validate on 15% (1041), and test on 15% (1041) of observations. The algorithm also imputed missing numbers with the median from each data category. The author used two evaluation methods: (1) plots of predicted versus observed linear fits; and, (2) a pseudo-R-squared measure calculated as the square root of the correlation between the predicted and observed values. He compared results from a random forest of 9,500 regression trees, and against results from a single regression tree (with 7 and 20 as the minimum and the maximum number of observations per split), a generalized linear model, and a neural network model. The author evaluated the pseudo-R-squared measure results twice, each using the validation and testing hold-back data sets that were randomly selected during partitioning and used the average of the two accuracy findings for the results. Minitab 19 (version 19.2020.1, Minitab LLC) was used to calculate means, medians, and 95% confidence intervals.

## Results

Based on the artificial intelligence and statistical analysis, 13 independent variables, each demonstrating a statistically significant relationship with incidence rate by a P-value < .05, explains 97.4% of the variability between incidence rates during the growth phase of the SARS-CoV-2 epidemic cycle across 88 days in 157 countries. Moreover, the algorithm predicts incidence rate per 100,000 with an average pseudo-R2 accuracy of 98.5% (validation = 98.7%, test = 98.2%; see Figure 1). The mean of squared residuals was 229.7, making the mean residual +/- 15.2 per 100,000. The mean incidence rate per 100,000 was 25.4 (95% CI 23.4 to 27.5), with a standard deviation of 86.7 (95% CI 85.3 to 88.2). An Anderson-Darling normality test indicated the data does not follow a specific, or normal, distribution (P-value < .005).

**Figure 1:**
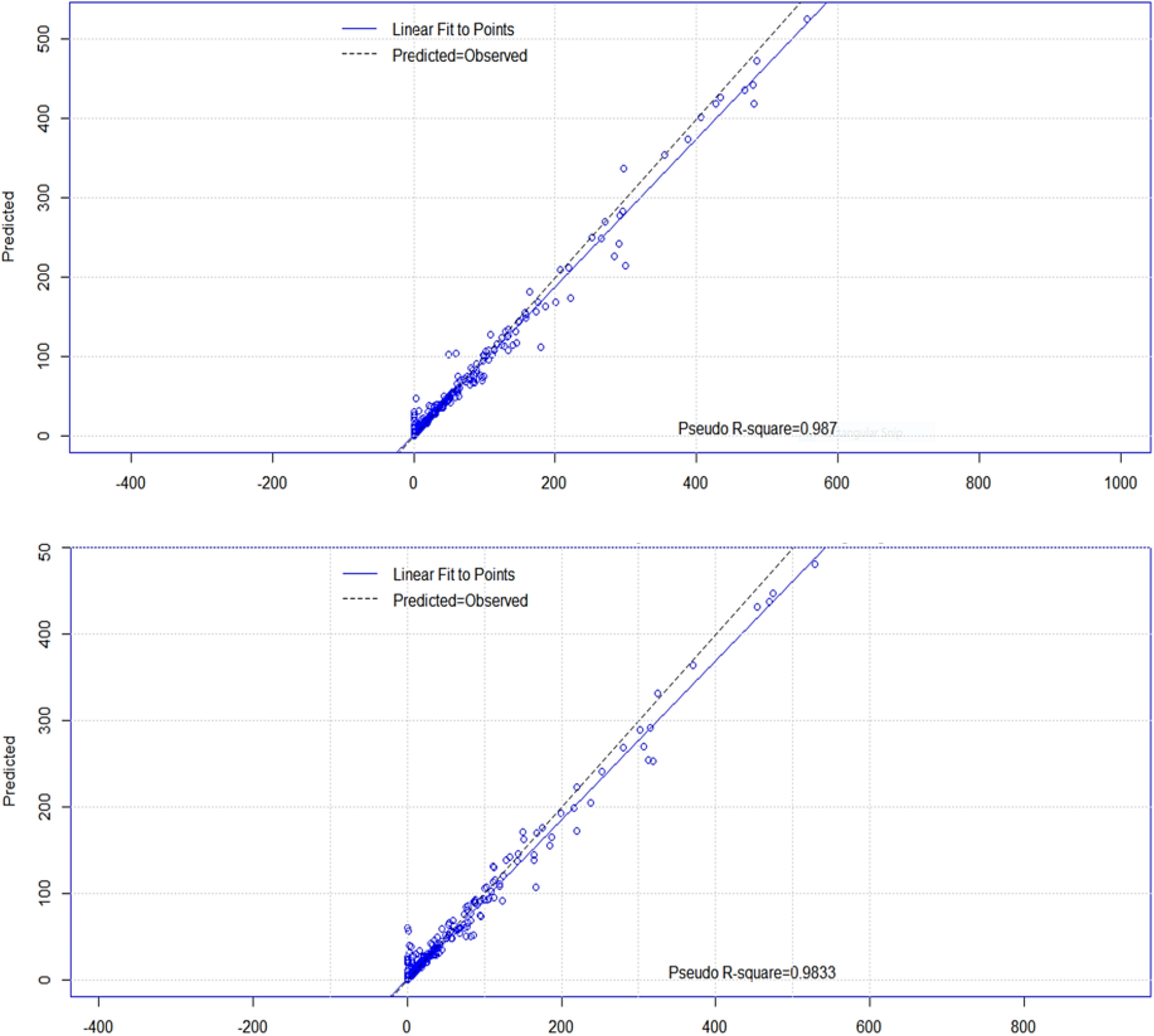
Fit of predicted vs. observed linear fit of SARS-CoV-2 incidence rate per 100,000 and pseudo-R2 of 15% validation hold-back data set (top) and 15% of test hold-back data set (bottom)

The relative importance of independent predictor variables was computed by percent increase in mean squared error (see Figure 2). The mean error is the average distance between the predicted and observed values. It is squared to ensure positive values and to weight greater distances. The percent increase in mean squared error is the proportional increase in the error of predictions when a variable is randomly excluded, or muted. For example, when the number of days since the index case was muted, the mean squared error increased by 212.4%, making it the most comparatively significant input predictor. Scores of tests of statistical significance, Spearman rho correlation strengths and directions, 95% confidence intervals, and high-level interpretations are in Figure 3.

**Figure 2:**
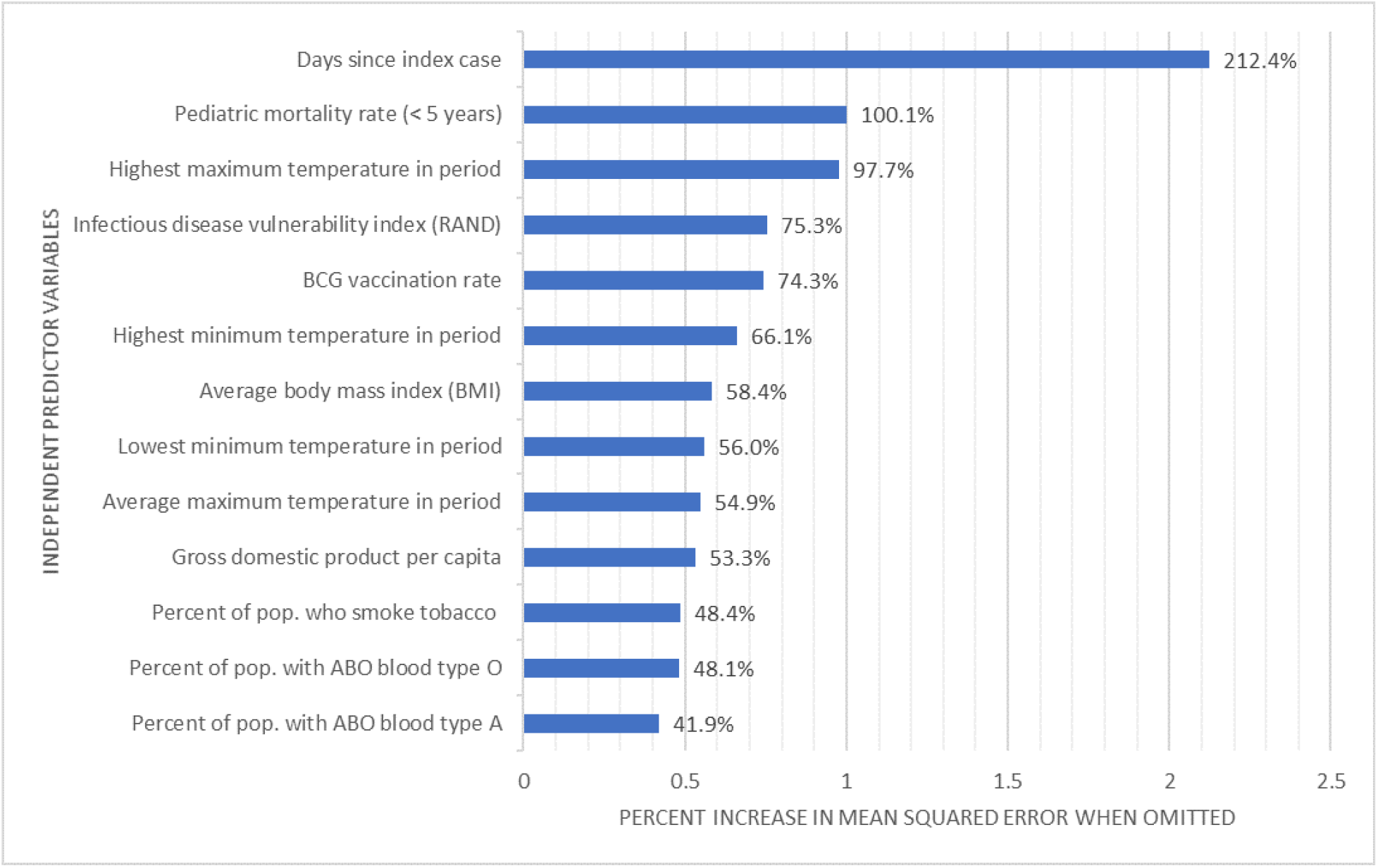
Comparative importance of independent predictors ranked by percent increase in mean squared error from exclusion

**Figure 3:**
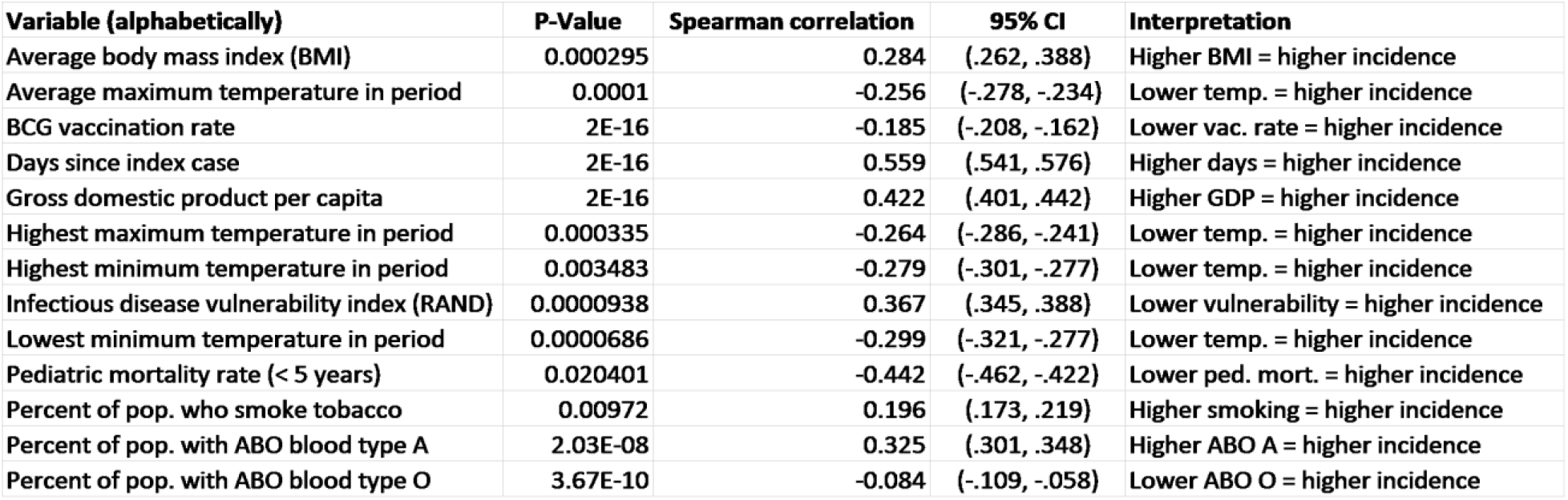
Table of independent predictor variable scores of probabilities of statistical insignificance, strength of correlation, confidence intervals, and interpretations

## Discussion

The primary importance of this work is a new capability to know in advance the order of magnitude of the disease burden of SARS-CoV-2 for any given population and time in a growth cycle. This new capability will enable leaders to make more accurate and precisely targeted decisions regarding public health interventions to minimize morbidity and mortality. For example, the incidence rates of those with a high BMI, smoke tobacco and have ABO blood type A are in three elevated risk groups for infection. (*5*)

The secondary importance of this work is new knowledge quantifying the relative importance of the social determinants that are root causes of incidence variance. This knowledge will enable leaders to target and prioritize programming more accurately. It is distinctly crucial because several of the findings are atypical from historic viral modulators. For example, to reduce SARS-CoV-2 morbidity and mortality, leaders may want to prioritize public health interventions focused on reducing body mass index, smoking, and pediatric mortality contributors because traditional infectious disease vulnerability and economic strength have a negative association with incidence rates. (*6*) Moreover, humidity and ultraviolet light exposure, which previous research suggests modulate the virus, have no statistically significant relationship with SARS-CoV-2 incidence variances.

The tertiary importance of this report is proof that machine learning methodologies can accurately and quickly inform our understanding of zoonoses’ disease dynamics with pandemic potential. For example, by entering dozens of possible demographic, economic, environmental, and public health measurements as independent predictors into machine learning algorithms, they can accurately determine within hours or days which factors explain inequalities in incidence, prevalence, or disease transmission. Moreover, the algorithms can also quantify and ordinally rank the social determinants to the root causes of variances.

This report has several limitations related to data dependencies of the model. One, because the current pandemic was seeded first and most heavily in more developed countries, it may have contributed to paradoxical findings such as higher incidence where infectious disease vulnerability is lower, and economies are more robust. Two, in geographically large countries, environmental measurements vary widely. Three, approximately 3,557 (3.7%) of 97,174 data points were missing and imputed with a median; actual observations may differ from the categorical medians. Four, the analysis was conducted mid-pandemic across only 88 days. Findings after the pandemic across its duration will be more definitive. Five, because testing availability was scant during the period of observation, the incidence rates measured probably reflect more severe cases that were symptomatic and hospitalized for testing rather than the actual incidence rate. This limitation could be significant if a large portion of those infected are asymptomatic but still contagious.

One implication of these findings is the importance of basic public health behaviors such as weight control and tobacco use, and the factors that contribute to pediatric survivability (e.g., education, nutrition, vaccinations). The second implication of these findings is that while previous research indicates viruses are modulated by temperature and humidity, these factors may only nominally slow the transmission of more contagious viruses. A third implication of these findings is that the causes of disease incidence variances are complex and sometimes surprising. A fourth and final implication of these findings is that the usefulness of machine learning as a public health tool is encouraging.

## Data Availability

The data set on which this study was based is available from the author upon request.

## Acknowledgements

*The author wishes to thank Dr. Luka Fajs and Mr. W. Andy Chang for their helpful comments on drafts of this article*.

## Notes

### Competing Interest Statement

The authors have declared no competing interest.

### Funding Statement

No external funding.

### Author Declarations

No IRB or ethics committee approvals were necessary because the study used secondary data from the public domain.

